# Pulsatile Gaussian-Enveloped Tones (GET) Vocoders for Cochlear-Implant Simulation

**DOI:** 10.1101/2022.02.21.22270929

**Authors:** Qinglin Meng, Huali Zhou, Thomas Lu, Fan-Gang Zeng

## Abstract

Acoustic simulations of cochlear implants (CIs) allow for studies of perceptual performance with minimized effects of large CI individual variability. Different from conventional simulations using continuous sinusoidal or noise carriers, the present study employs pulsatile Gaussian-enveloped tones (GETs) to simulate several key features in modern CIs. Subject to the time-frequency uncertainty principle, the GET has a well-defined tradeoff between its duration and bandwidth. Two types of GET vocoders were implemented and evaluated in normal-hearing listeners. In the first implementation, constant 100-Hz GETs were used to minimize within-channel temporal overlap while different GET durations were used to simulate electric channel interaction. This GET vocoder could produce vowel and consonant recognition similar to actual CI performance. In the second implementation, 900-Hz/channel pulse trains were directly mapped to 900-Hz GET trains to simulate the maxima selection and amplitude compression of a widely-used *n*-of-m processing strategy, or the Advanced Combination Encoder. The simulated and actual implant performance of speech-in-noise recognition was similar in terms of the overall trend, absolute mean scores, and standard deviations. The present results suggest that the pulsatile GET vocoders can be used as alternative vocoders to simultaneously simulate several key CI processing features and result in similar speech perception performance to that with modern CIs.

## I. Introduction

VOVODERs as a means of speech synthesis have a long and rich history. At the 1939 New York World’s Fair, Homer Dudley of Bell Labs demonstrated his vocoder invention that could “remake speech” automatically and instantaneously (18-ms delay) by controlling energy in 10 frequency bands (from 0 to 3000 Hz) that contained either buzz-like tone or hiss-like noise carriers [1]. He later realized that the vocoder could be used in synthesizing speech, and transformed in various ways to study the relative contributions of fundamental parameters in speech synthesis and recognition. He found that good intelligibility can be achieved by controlling “only low syllabic frequencies of the order of 10 cycles per second”, whereas the emotional content of speech can be controlled by altering the frequency of the buzzing tones.

The early multi-channel CIs followed Dudley’s original vocoder idea closely by extracting and delivering speech fundamental frequency (F0) in the form of electric pulse rate and one or two formants (F2 or F1/F2) in the form of electrode position [2, 3]. The speech understanding of the early CIs was relatively low (<50% correct for sentence recognition in quiet), due not only to crude F0 and formant extraction methods (i.e., zero-crossing) at that time, but, more importantly, to the complicated interactions between sound frequency and electric pitch, for example, individual variability in electrode insertion angle or depth, cochlear vs. ganglion cell tonotopic organization, current spread, and nerve survival. These interactions make accurate F0 and formant representation difficult if not impossible even if both F0 and formants can be exactly extracted by today’s algorithms. As a result, contemporary CIs have abandoned the F0 and formant extraction method but adopted speech processing strategies that extract band-specific temporal envelopes from 8-24 frequency bands. The envelopes are used to amplitude modulate a continuous, but fixed, high-rate (at least two to four times the highest envelope frequency) pulse train, which is then delivered to a corresponding electrode in an interleaved fashion in which no two electrodes fire simultaneously [4, 5]. These advances in multi-channel CIs have produced 70-80% correct sentence recognition in quiet, which is sufficient for an average user to carry on a conversation without lipreading [6].

Acoustic simulations of CIs have been developed and widely used [7] for at least three reasons. First, acoustic simulations minimize the effect of large CI individual variability (e.g., cognitive differences, demographic variables, and electrode-neuron interface), which may confound or mask the relative importance of speech processing parameters, e.g., [5]. Second, acoustic simulations allow the evaluation of relative contributions of different cues to auditory and speech perception, e.g., [8] and [9]. Third, acoustic simulations allow a normal-hearing listener to appreciate the quality of CI processing and the degree of difficulty facing a typical CI user.

Traditionally, acoustic simulations of CIs have used either noise- [10] or sinusoid-excited [11] vocoders. In these vocoders, the noise or sinusoid simulates the electric pulse train, while the number of frequency bands and their overlaps simulate the limited number of electrodes and their current spread, e.g., [10]. A significant drawback of these traditional vocoder models is the lack of simulation of the pulsatile nature of CI electric stimulation. Several studies have attempted to develop acoustic models that simulate pulsatile electric stimulation, such as filtered noise bursts [12, 13], filtered harmonic complex tones [14], and pulse-spread harmonic complexes [15, 16]. However, there are limitations to those methods in simulating some important features in modern CIs. First, these vocoders cannot simulate the discrete nature of pulsatile stimulation on a pulse-by-pulse basis. Second, they do not allow independent manipulation of the overlap between spectral and temporal representation. Third, it is difficult for vocoders using continuous carriers to simulate some CI speech processing strategies, e.g., *n*-of-*m*, in which the low-energy bands are abandoned to produce temporally separated envelopes.

Here we identified the Gabor atom [17], also known as the Gaussian-enveloped tone (GET), as a means of simulating the essential features of modern CI processing as discussed above. The GET has been used to study a wide range of auditory phenomena in normal-hearing or hearing-impaired listeners, e.g., temporal gap detection [18, 19], intensity discrimination [20-23], simultaneous and non-simultaneous masking [24, 25], interaural timing difference (ITD) [26], and cortical encoding of pulsatile stimulation [27-29]. More recently, GET train has been used to simulate some basic tasks on binaural hearing with CIs, e.g., sound localization [30, 31], lateralization [32], binaural masking level differences [33], temporal weighting of ITD and interaural level difference (ILD) [34], effects of electrode place mismatch on binaural cues [35, 36], and effects of temporal quantization on ITD discrimination [37].

In signal processing, due to the time-frequency uncertainty principle (also referred to as the Gabor limit), the duration and bandwidth of a signal cannot be independently controlled, and their product is no lower than a limit, which is reachable only by GETs (or say Gabor atoms) [17, 38, 39]. This is an important reason why most of the above-mentioned psychoacoustic studies use GETs as stimuli.

However, the performance of GET-based vocoders in simulating speech perception with CIs has not been investigated. In much of the existing literature, conventional channel-vocoders with eight channels using continuous noise or sine-wave carriers were used to replicate the sound of 12-24 channel CIs. The main reason is the performance of eight-channel vocoders in normal-hearing listeners usually matches the better performance of actual CI users [40].

This study introduces a novel GET vocoder and demonstrates its potential for simulating CI speech perception. In the following sections, the implementation and theory of the proposed GET vocoders are introduced in detail; then two separate experiments of speech perception, each with a different type of GET vocoder, are used to demonstrate the potential of the novel pulsatile vocoders on CI speech perception simulation. Specifically, the first GET [30, 41] is a naïve type using non - interleaved 100-pps (pulse per second) GET trains as carriers to study the effect of current interaction among channels. The second GET [42, 43] is an advanced type that can directly map individual electric pulses from a clinical n-of-m strategy with 900-pps pulse rate into an acoustic GET. In this way, any CI electrodogram (not limited to the selected strategy) can be directly transformed into a vocoded sound. Such direct transformation can simulate not only pulsatile timing cues but also many other features of CI electric stimuli (e.g., amplitude compression and maxima selection).

The pulsatile GET vocoder can replicate the temporal (pulsatile), intensity (compressed and quantized), and spectral (maxima-selected) features of an actual CI strategy. Furthermore, current spread at individual electrodes can be simulated by changing the GET bandwidth through the pulse duration parameter. We hypothesized that the GET vocoder could be an alternative vocoder model to simulate speech perception with CIs. Nevertheless, the uncertainty principle imposes unavoidable physical constraints on the time-frequency tradeoff, which might limit the performance of the pulsatile simulation and should be carefully controlled.

## II. GET Theory And Vocoder Algorithms

### A. GET Theory

A Gaussian function is symmetrical in the time domain:

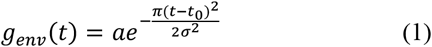

where *a* determines the function’s maximum amplitude, *t*_0_ the maximum amplitude’s temporal position, and *σ* the effective duration or 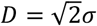, at which the amplitude is 6.82-dB down from the maximum amplitude [20]. Its Fourier transform is:

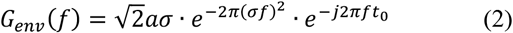

The shape of its amplitude spectrum is also a Gaussian function with an effective bandwidth between the 6.82-dB down cutoff frequencies.

The effective duration (D) and the effective bandwidth (B) can be traded:

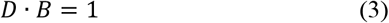

meaning that increasing the duration will narrow the bandwidth and vice versa.

Acoustic simulation of a single electric pulse in a frequency channel can be generated by multiplying the above Gaussian function by a sinusoidal carrier:

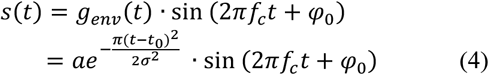

where s(t) has the same effective duration and effective bandwidth as *g*_*env*_(*t*) except for changing the center frequency from 0 to *f*_*c*_, and *φ*_0_ is an initial phase.

Fig. 1 illustrates both waveform (a) and spectrum (b) of a unit-amplitude Gaussian-enveloped single pulse (i.e., *a* = 1 in (4)). The carrier frequency *f*_*c*_ is 5 kHz. The 6.82-dB cutoff point (corresponding to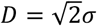) with an amplitude of 0.456 in Fig.1 was derived by substituting 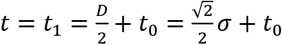 into (1), i.e.,

**Fig. 1.**
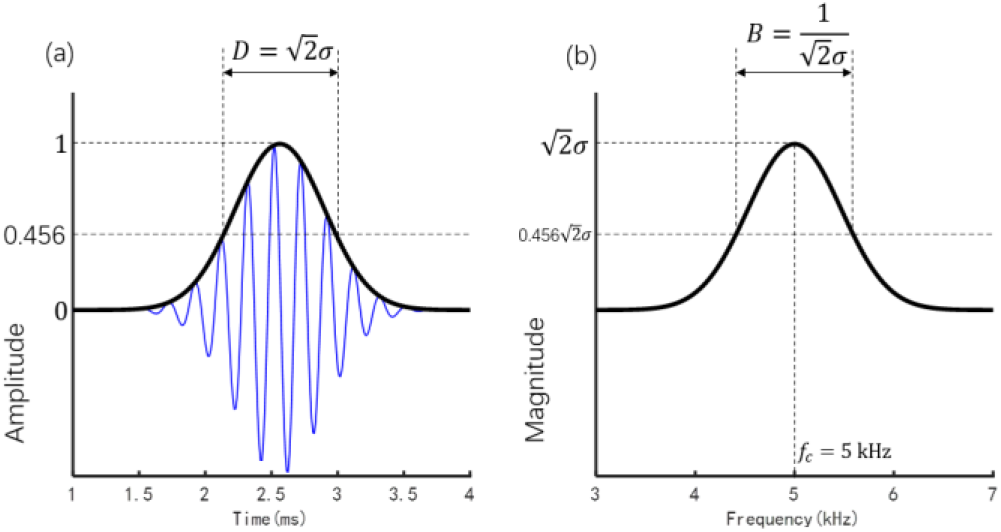
A unit-amplitude single pulse with Gaussian-shaped envelope (black line) in both the time (a) and frequency (b) domains. The carrier frequency is 5 kHz (the blue waveform in the left panel and the frequency with maximum amplitude in the right panel). The *σ* equals to 3/*f*_*c*_ = 0.6 ms in (1), producing an effective duration of 0.85 ms and an effective bandwidth of 1.2 kHz.

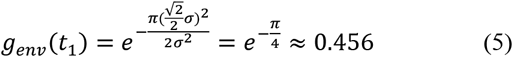

Using the GET defined by (4), the change of amplitude and timing of an electric pulse can be simulated by manipulating *a* and *t*_0_ respectively. Acoustic simulation of a continuous electric pulse train can be constructed by periodically repeating *s*(*t*) or convolution of the electric pulse train and a GET.

Different from the CI electric pulses with constant duration at the order of tens of microseconds, the GET duration should be much longer to contain at least several (*l*) periods (e.g.,*l* = 2, 3, *or* 4) of the tone carrier. Therefore, the carrier period or frequency will determine the lower limits of the GET duration. The three lines in the two panels of Fig. 2 illustrate the dependent relationship between the GET duration (bandwidth), pulse rate, and carrier frequency, when 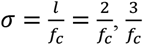, and 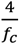, respectively. The GET effective bandwidth equals in value to the maximum pulse rate that can be transmitted without obvious temporal interaction between neighboring GETs. Here the GET duration threshold for the “obvious temporal interaction” was defined as the effective duration of GET, i.e., 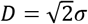. Increasing the duration (i.e., larger σ) can decrease the bandwidth with the maximum rate decreasing correspondingly.

**Fig. 2.**
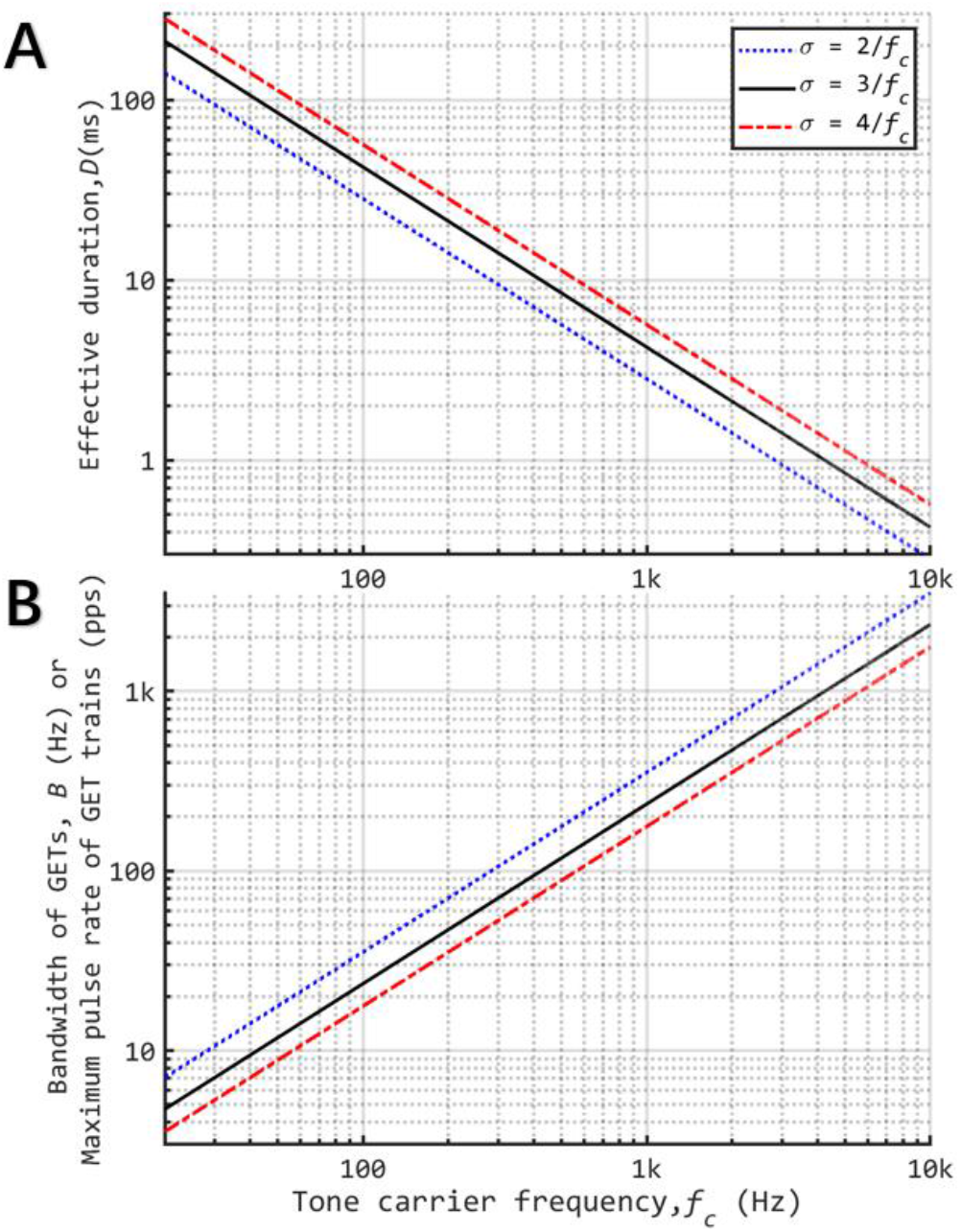
The relationship between the tone carrier frequency and the effective duration 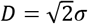 (see Panel A) or effective bandwidth *B* = 1/*D* (see Panel B) of Gaussian-enveloped tones (GETs). All axes are logarithmically scaled. The *σ* was assumed to be 2/*f*_*c*_, 3/*f*_*c*_, or 4/*f*_*c*_ to demonstrate the effects of different duration of GETs. For certain combinations of *f*_*c*_ and σ, the maximum GET rate that can be transmitted with no temporal interaction between neighboring GETs is 1/*D*, which equals in value to the effective bandwidth in Panel B.

At frequency bands with high carrier frequencies above ∼2.5 kHz (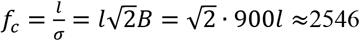, 3818, and 5091 Hz for *l* = 2, 3, and 4, respectively), a conventional pulse rate of 900 pps could be simulated without obvious temporal interaction between neighboring GETs. For carrier frequencies within the middle-frequency range around 2 kHz, the 900 pps is still possible to simulate, but neighboring GETs have moderate temporal interaction. The amplitude of the crossing point of neighboring GETs at a 2 kHz carrier would be

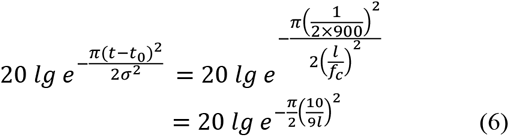

whose values are −4.21, −1.87, and −1.05 dB (relative to the maximum amplitude) for 2, 3, and 4, respectively. For a low-frequency carrier, the pulsatile feature for simulation of individual electric pulses cannot be guaranteed due to temporal interactions between neighboring GETs.

The temporal envelopes delivered in electric speech stimuli are often temporally separated across channels in many CI strategies, as speech contains natural gaps within each channel of signal between syllables, and frame-wise low power bands are temporarily abandoned resulting from the maxima selection for *n*-of-*m* strategies. Additionally, envelope energies lower than the compression threshold level (or T level) are not represented in electric stimuli (i.e., no stimulation) in some strategies. For the temporally separated electric stimuli within each channel, GET carriers can better represent temporal separation features as well as CI compression (limited electric dynamic range), both of which are often omitted in conventional noise and sine-wave vocoders. The temporal separation features may be simulated in all channels, and the low carrier frequency limit *f*_*c*_*low*_ is mainly determined by the duration *d*_*gap*_ of each gap in the pulse trains:

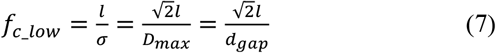

where *D*_*max*_ is the maximum possible GET duration, which equals the gap duration.

Current (or spectral) spread was acknowledged to be an important issue influencing the frequency resolution of CIs [44]. For a single GET (defined by (4)), its bandwidth is determined by its duration due to the time-frequency uncertainty principle. Therefore, it is possible to simulate CI current spread by manipulating the GET duration, meaning the pulsatile timing feature and the current spread cannot be independently manipulated.

In short, the GETs can simulate and manipulate five important parameters of CI processing or stimulation: (1) pulse rate by changing the period of pulse generation, (2) temporal envelope (including its compression and quantization) by changing the amplitude of individual GETs in a pulse train within a channel, (3) spectral envelope by changing the GET amplitude across channels, (4) place of excitation by changing the carrier tone frequency, and (5) spread of excitation by changing the effective bandwidth in GETs. The precise manipulation of these five important parameters allows acoustic simulation of modern CIs using pulsatile electric stimulation. The limitations from the dependent relationships between duration, bandwidth, and carrier frequency of GETs are discussed above and should be taken into consideration during algorithm design and experiments of CI simulations with GETs.

### B. Vocoder Algorithm Frameworks

Fig. 3A shows the conventional acoustic simulation of CI using either noise [10] or sine-wave vocoders [11]. The output filters can be used to control the current spread, but no temporal separation feature (e.g., pulsatile timing and temporally separated envelope) can be simulated.

**Fig. 3.**
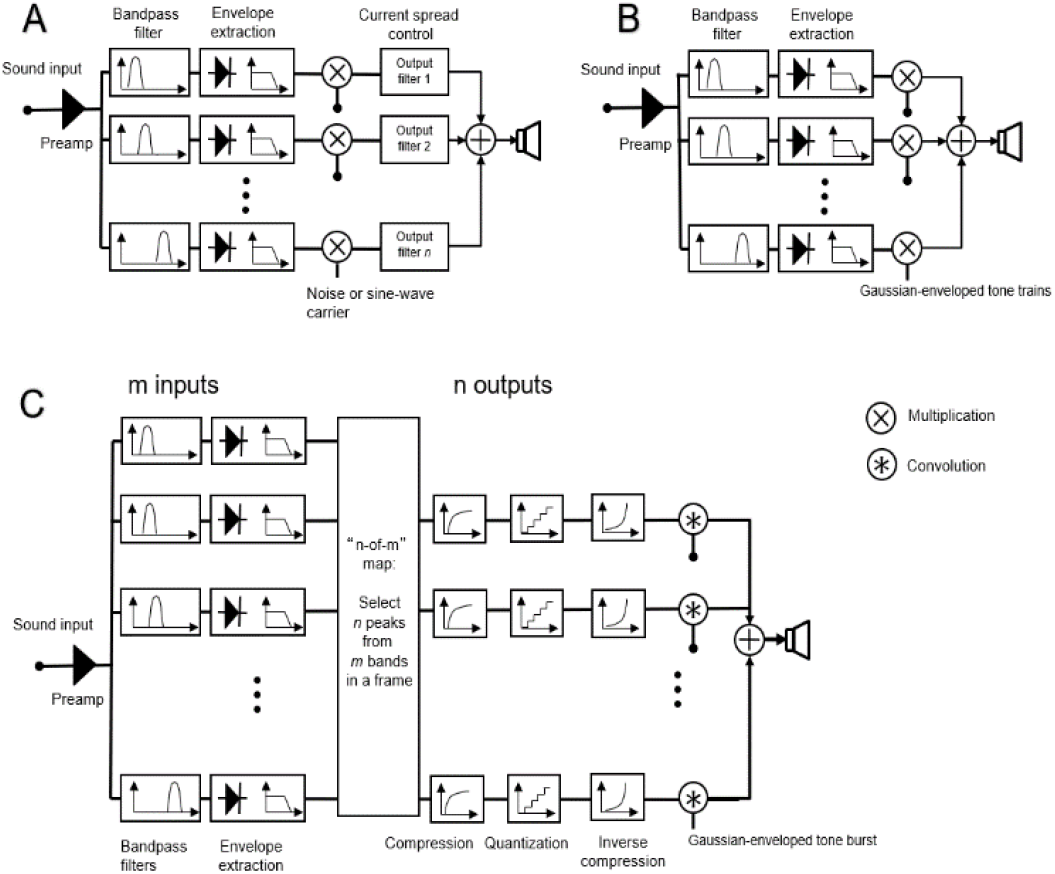
Block diagrams of conventional channel vocoder (A), the first (B) and second (C) types of GET vocoders. The pulsatile vocoders are using GETs as carriers (the first type; used in Exp. 1) or using a single GET as an impulse response (the second type; used in Exp. 2). The front-end pre-emphasis, bandpass filter, and envelope extraction can be implemented either in the temporal or spectral domain.

The first GET vocoder was proposed by [41] (see Fig. 3B) and subsequently used in a sound localization study [30]. As a naïve implementation, this approach replaces the conventional continuous carriers with pulsatile GET carriers. To demonstrate the effects of current interaction realized by different GET durations, vowel and consonant perception with non-interleaved 100-pps GET carriers was measured in Experiment 1 (Section III).

The second GET vocoder was proposed by [42] (see Fig. 3C). Compared to the naïve implementation of the first type, the second GET vocoder hypothesized that a direct mapping from individual CI electric pulses to individual GET acoustic pulses could transmit similar speech information in both stimuli of CI and GET simulation. The implementation framework of the second GET vocoder considers a common feature of temporal-frame-based *n*-of-*m* selection in some CI processing strategies. The *n*-of-*m* selection means *n* maximum envelope values are selected out of the envelope values from the m input channels within a given time window. In this framework, the amplitude compression and quantization widely used in modern CIs can also be simulated. In Experiment 2 (Section IV), sentence intelligibility tests were carried out to demonstrate the feasibility of GET simulation on speech perception with the advanced combination encoder (ACE) strategy, which is a typical *n*-of-*m* strategy and has a default pulse rate of 900 pps.

The front-end processing stages of the three methods in Fig.3 share the same blocks of band-pass filters and envelope extraction, e.g., in a traditional temporal envelope-based continuous interleaved sampling (CIS) [4] or ACE strategy [45]. Details about the implementations of the two types of GET vocoders are provided in the following two experiment sections.

## III. Experiment 1: Simulation Of Current Spread

### A. Rationale

Experiment 1 was designed to study vowel and consonant speech perception with the first type of GET vocoder [30, 41] using non-interleaved GET carriers (where the GET centers for all channels are in alignment with each other in each frame). The interleaved sampling feature of modern CI strategies was not considered. A low pulse rate of 100 pps, which is much lower than the standard clinical rate (e.g., 900 pps or faster), was used in this experiment to minimize the within-channel inter-pulse temporal interaction. The primary purpose of this experiment is to examine the effects of current spread stimulated by manipulating the GET duration based on the uncertainty principle.

There is a substantial difference in simulating the spread of excitation between the conventional vocoder [10, 11] and the GET implementation [41]. In the conventional simulation, the spread of excitation is manipulated by changing the filter type, the bandwidth of the noise carriers, or the bandwidth of the synthesis band-pass filters at the vocoder output stage [46]. For the GETs, the spread of excitation is manipulated by increasing or decreasing the Gaussian tone duration, which produces a corresponding change in narrowing or widening the spectral bandwidth for each pulse.

### B. Methods

Five vocoders were used: three conventional vocoders - sine-wave, noise-separate, and noise-spread (Fig. 3A) - and two proposed vocoders incorporating the GET simulation -GET-separate and GET-spread (Fig. 3B).

Analysis processing of all five vocoders: The analysis filter banks consist of *M* band-pass filters (4th order Butterworth). The frequency spacing for cutoffs for the filter bank was defined in the range of [80, 7999] Hz according to a Greenwood map [47] (See Tab. I). The filtered signals were half-wave rectified and low-pass filtered (50 Hz 4th order Butterworth) to extract the envelope for each channel.

Synthesis processing for the conventional vocoders: For the sine-wave vocoder, a sine wave with a frequency centered at the corresponding analysis filtering band was used as the carrier. For the noise-separate vocoder, band-pass noise carriers were generated by passing white noise through filters that were the same as the analysis filters. The noise-separate vocoder, which results in minimal overlap between channels, should results in high speech perception accuracy. For the noise-spread vocoder, low-pass filters (4^th^ order Butterworth) were used to pass white noise for generating low-pass noise carriers. The cutoff frequencies of the low-pass filters were the same as the upper cutoff frequencies of the analysis filters. Low-pass filters were chosen to represent severe interactions between channels (especially on the low-frequency side), and provide a lower bound of performance with simple manipulation. For the two noise vocoders, after modulating each channel of filtered noise with the channel envelope, the output was filtered again to band-limit each channel. The band-limiting filters are the same as those used for the noise carrier generation. The final vocoded signal was synthesized by summing all channels.

#### Synthesis processing for the GET vocoders

For the GET vocoders, instead of modulating a filtered noise signal at the synthesis stage, the envelope in each channel modulates the amplitude of a GET train. Fig. 4 shows a 100-Hz pulse train, repeating the single pulse every 10 ms. The pulse train’s spectral envelope remains the same as the single pulse but its spectral fine structure becomes discrete with 100-Hz spacing (in this case, the maximum-amplitude frequency is 5 kHz with symmetrically decreasing-amplitude components at 4.9, 4.8, 4.7… and 5.1, 5.2, 5.3… kHz, respectively, see inset in the right panel). For the GET-separate vocoder, 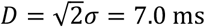, while for the GET-spread vocoder, 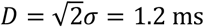. Because the first experiment focused on the spread of excitation, the pulses among all channels were synchronized, meaning that the “interleaved sampling” feature was not simulated.

**Fig. 4.**
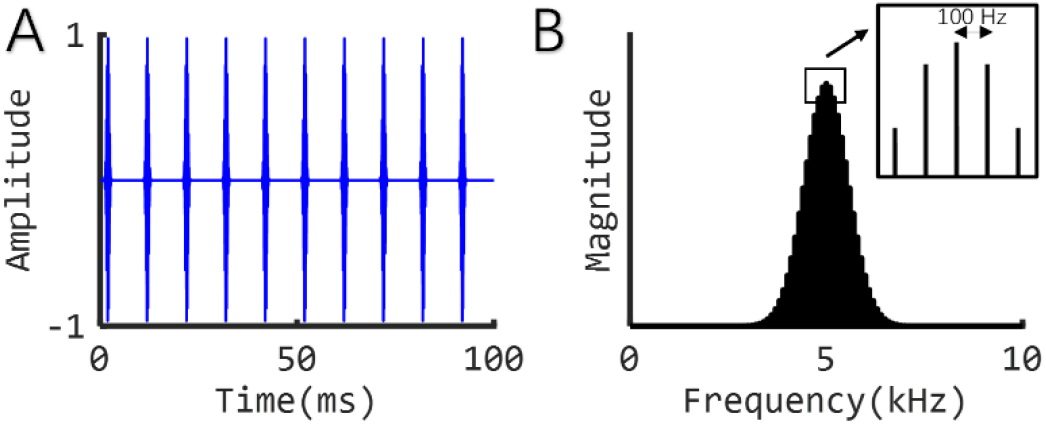
A 100-Hz pulse train, repeating a single pulse every 10 ms, in both the time (left panel) and frequency (right panel) domains. The parameters of the individual pulses are the same as those in Fig. 1.

CI stimulation was simulated using the above five different vocoders, i.e., sine-wave, noise-separate, noise-spread, GET-separate, and GET-spread. The numbers of channels tested were 2, 4, 8, 16, and 32. There were 12 medial vowels and 14 medial consonants in the vowel and consonant tests, respectively. Fig. 5 provides an example of 16-channel vocoded stimuli for vowel tests. Each stimulus was presented 10 times. Stimuli were presented through headphones (HDA 200, Sennheiser), and the sound level was calibrated to 70 dB SPL. This procedure was conducted following procedures approved by the University of California Irvine Institutional Review Board.

**Fig. 5.**
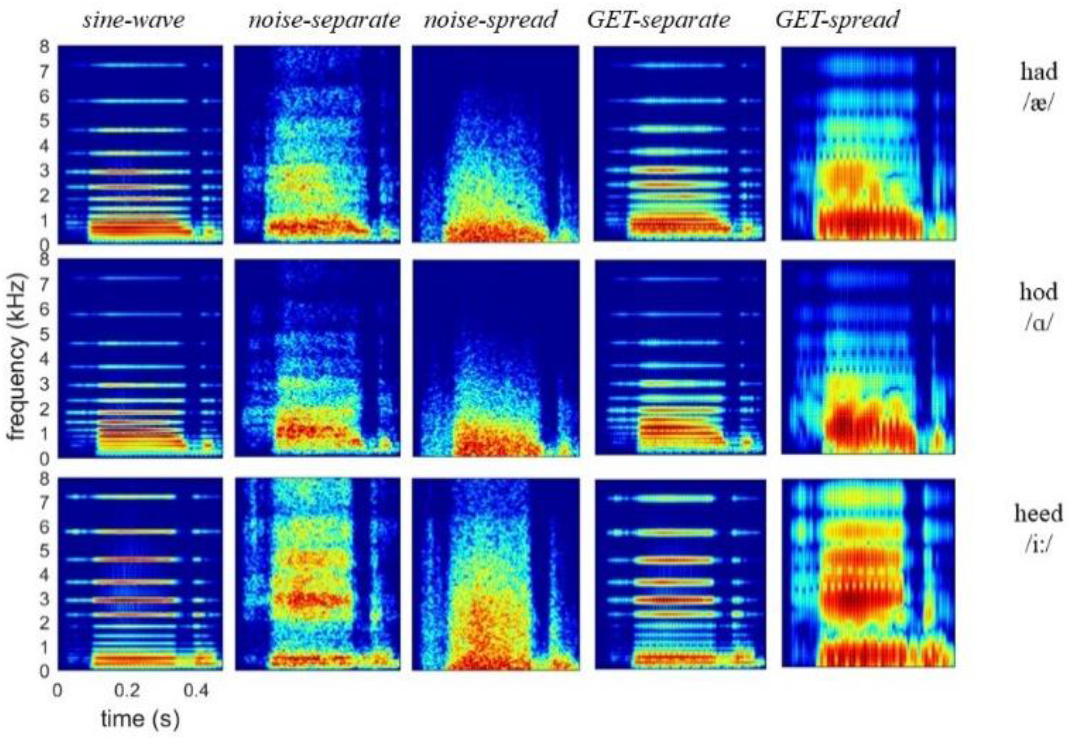
Spectrograms of three vowel stimuli encoded by the sine-wave, noise-separate, noise-spread, GET-separate, and GET-spread vocoders with 16 channels.

Seven normal hearing (NH) participants, ages 18-21, were tested in an anechoic chamber (IAC) using the English vowel and consonant recognition tests adopted from [48].

### C. Results

Results are shown in Fig. 6. For the vowel test, the seven NH participants scored approximately 20% under all simulation conditions with two channels. Increasing the number of channels also improved performance. With eight channels, performance under the different conditions began to separate. The sine-wave vocoder outperformed actual CI data, which showed no improvement beyond 8 channels. The noise-separate vocoder and GET-separate vocoder showed similar performance trends. When electrode interaction was simulated with overlapping filters, the subject performance showed a plateau near 60% with noise-spread, similar to actual CIs. The GET-spread condition underperformed CI data in this case, saturating near 35% with eight channels.

**Fig. 6.**
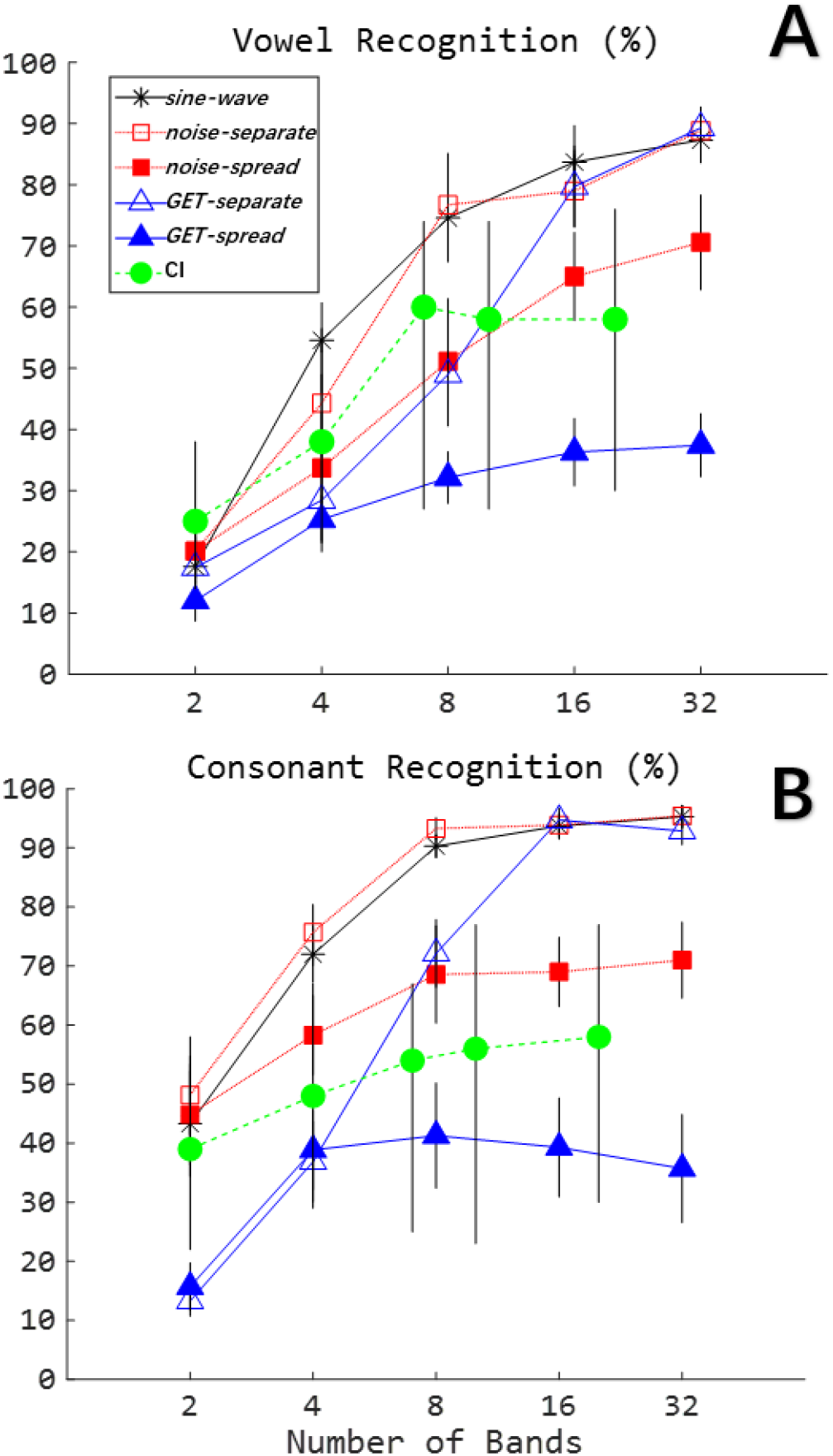
Vowel (A) and consonant (B) recognition as a function of number of bands (channels). Simulation data are averaged from seven normal hearing subjects listening to vocoded speech. For the simulation data, standard errors are indicated by the vertical bars. For the CI data, the bars show the entire ranges of performance across all their 19 participants.

Further, a two-way repeated-measures ANOVA with Geisser-Greenhouse correction was used to analyze the vowel simulation results with vocoder and number of bands as the main factors. The effect of vocoder (*F*_1.987, 11.92_ = 49.87, *p* < 0.0001), number of bands (*F*_2.018, 12.11_ = 90.66, *p* < 0.0001), and their interaction (*F*_3.890, 23.34_ = 9.842, *p* < 0.0001) were all significant. To further analyze these effects, multiple comparisons with Bonferroni corrections were implemented for each vocoder (to compare the five band numbers) and for each band number (to compare the five vocoders). Table II shows the results of multiple comparisons between different numbers of bands for each vocoder. Generally, there was a trend of better performance with more bands. Still, the mean scores were not significantly different for 8, 16, and 32 bands (the only exception was 8 vs. 32 with GET-separate). Table III shows the results of multiple comparisons between vocoders for each number of bands. Because at 2 and 4 bands most vocoder pairs showed no significant mean difference (the only exception was sine-wave vs. noise-spread at 4 number of bands with p = 0.009), the comparison results were not listed. GET-spread derived the lowest scores among the five vocoders at 16 and 32 bands, while GET-separate did not show significantly different mean scores from the other three vocoders. The sine-wave, noise-separate, and GET-separate vocoders did not show significantly different mean scores.

**TABLE I.** Cutoff Frequencies of The Band-pass Filters in Exp.1 According to a Greenwood Map

| Number of bands (M) | Cutoff frequencies (Hz) |
| --- | --- |
| 2 | 80, 1250, and 7999 |
| 4 | 80, 424, 1250, 3234, and 7999 |
| 8 | 80, 215, 424, 748, 1250, 2028, 3234, 5103, and 7999 |
| 16 | 80, 140, 215, 308, 424, 568, 748, 972, 1250, 1597, 2028, 2565, 3234, 4067, 5103, 6393, and 7999 |
| 32 | 80, 108, 140, 176, 215, 259, 308, 363, 424, 492, 568, 653, 748, 854, 972, 1103, 1250, 1414, 1597, 1801, 2028, 2282, 2565, 2881, 3234, 3628, 4067, 4556, 5103, 5713, 6393, 7152, and 7999 |

**TABLE II.** Results of Multiple Comparisons Between Vowel Recognition Scores With Five Band Numbers for Each of The Five Vocoders.

| vocoder→<br>number of<br>bands pair↓ | sine-wave |  | noise-separate |  | noise-spread |  | GET-separate |  | GET-spread |  |
| --- | --- | --- | --- | --- | --- | --- | --- | --- | --- | --- |
|  | mean<br>diff | *p* | mean<br>diff | *p* | mean<br>diff | *p* | mean<br>diff | *p* | mean<br>diff | *p* |
| 2 vs. 4 | *37.0 | 0.014 | 24.1 | 0.140 | 13.6 | 1.000 | 11.0 | 0.827 | *13.3 | 0.019 |
| 2 vs. 8 | **57.0 | 0.001 | **56.6 | 0.002 | 31.0 | 0.085 | 31.6 | 0.082 | *20.1 | 0.048 |
| 2 vs. 16 | ***66.1 | <0.001 | ***58.9 | <0.001 | **44.9 | 0.001 | ***62.3 | <0.001 | *24.3 | 0.005 |
| 2 vs. 32 | ***69.7 | <0.001 | ***68.7 | <0.001 | **50.4 | 0.001 | ***71.9 | <0.001 | *25.4 | 0.012 |
| 4 vs. 8 | 20 | 0.076 | *32.4 | 0.019 | 17.4 | 0.057 | 20.6 | 0.209 | 6.9 | 0.564 |
| 4 vs. 16 | *29.1 | 0.049 | *34.7 | 0.023 | ***31.3 | <0.001 | **51.3 | 0.002 | *11.0 | 0.034 |
| 4 vs. 32 | *32.7 | 0.037 | *44.6 | 0.011 | ***36.9 | <0.001 | ***60.9 | <0.001 | 12.1 | 0.296 |
| 8 vs. 16 | 9.1 | 0.199 | 2.29 | 1.000 | 13.9 | 0.147 | 30.7 | 0.058 | 4.1 | 1.000 |
| 8 vs. 32 | 12.7 | 0.288 | 12.1 | 0.696 | 19.4 | 0.059 | *40.3 | 0.011 | 5.3 | 1.000 |
| 16 vs. 32 | 3.6 | 1.000 | 9.86 | 0.131 | 5.6 | 0.107 | 9.6 | 0.455 | 1.1 | 1.000 |

**TABLE III.** Results of Multiple Comparisons Between Vowel Recognition Scores With Five Vocoders for Each of The Three Band Numbers (8, 16, and 32).

| number of bands→<br>vocoder pair↓ | 8 |  | 16 |  | 32 |  |
| --- | --- | --- | --- | --- | --- | --- |
|  | mean diff | *p* | mean diff | *p* | mean diff | *p* |
| sine-wave vs. noise-separate | -2.1 | 1.000 | 4.7 | 1.000 | -1.6 | 1.000 |
| sine-wave vs. noise-spread | 23.4 | 0.054 | **18.7 | 0.009 | 16.7 | 0.120 |
| sine-wave vs. GET-separate | 25.6 | 0.065 | 4.0 | 1.000 | -2.0 | 1.000 |
| sine-wave vs. GET-spread | **42.4 | 0.002 | ***47.4 | <0.001 | ***49.9 | <0.001 |
| noise-separate vs. noise-spread | *25.6 | 0.030 | *14.0 | 0.025 | *18.3 | 0.048 |
| noise-separate vs. GET-separate | 27.7 | 0.080 | -0.7 | 1.000 | -0.4 | 1.000 |
| noise-separate vs. GET-spread | **44.6 | 0.004 | ***42.7 | <0.001 | ***51.4 | <0.001 |
| noise-spread vs. GET-separate | 2.1 | 1.000 | -14.7 | 0.130 | -18.7 | 0.102 |
| noise-spread vs. GET-spread | 19.0 | 0.459 | **28.7 | 0.002 | **33.1 | 0.004 |
| GET-separate vs. GET-spread | 16.9 | 0.156 | **43.4 | 0.001 | ***51.9 | <0.001 |

Consonant recognition showed similar performance trends across the simulation types, with sine-wave, noise-separate, and GET-separate outperforming CIs (adapted from [48]) when there were eight or more channels simulated. Noise-spread brought the performance closer to actual CI data, while again GET-spread resulted in worse performance than that observed with CI listeners. With only two channels, both GET-separate and GET-spread showed much lower performance than actual CIs. For the simulation results, consonant recognition scores were analyzed using the same statistical method as the above vowel data analysis. The effects of vocoder (*F*_1.404, 8.427_ = 62.55, *p* < 0.0001), number of bands (*F*_2.234, 13.40_ = 379.0, *p* < 0.0001), and their interaction (*F*_3.080, 18.48_ = 10.88, *p* = 0.0002) were all significant on consonant recognition. Results of multiple comparisons are shown in Table IV and V. The relative scores show similar trends as the results of multiple comparisons for vowel recognition (see Table II and III).

**TABLE IV.**
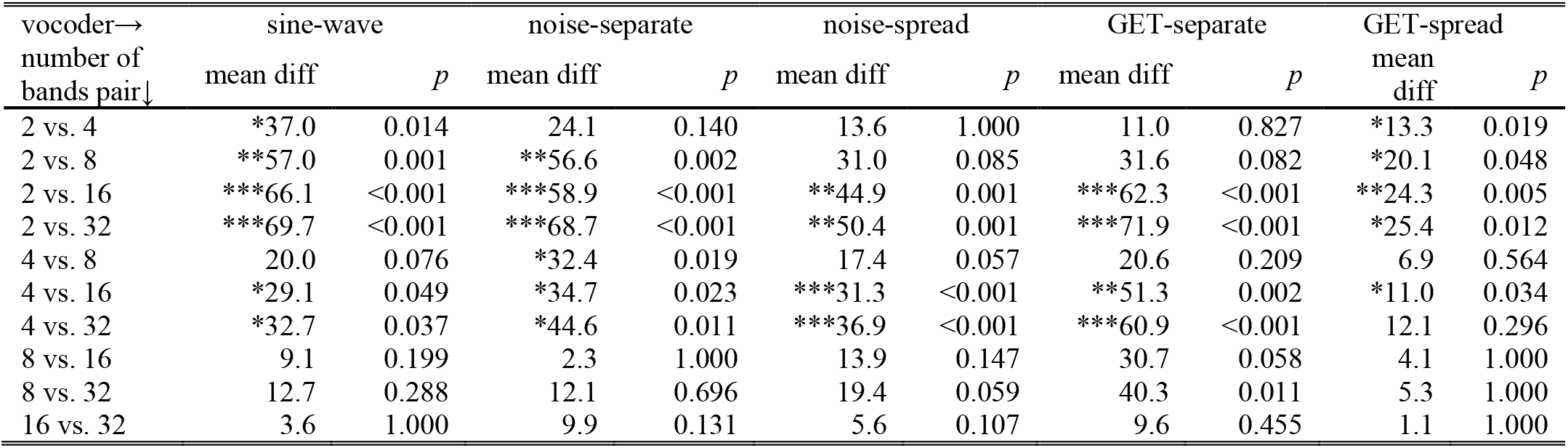
Results OF Multiple Comparisons Between Consonant Recognition Scores With Five Number of Bands for Each of The Five Vocoders

**TABLE V.**
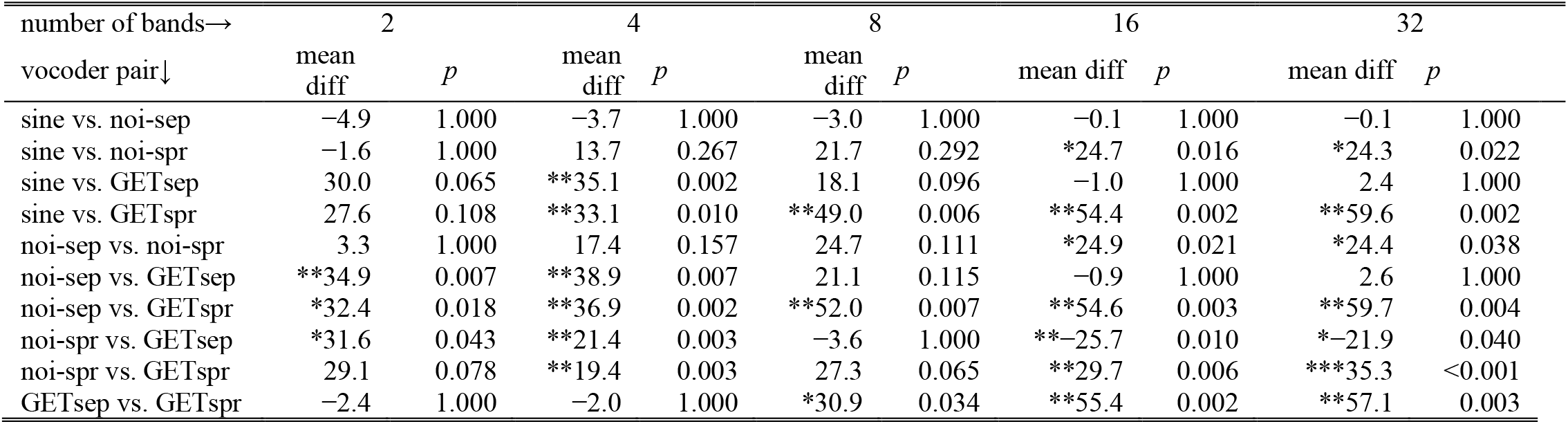
Results of Multiple Comparisons Between Consonant Recognition Scores With Five Vocoders for Each of The Five Band Numbers

**TABLE VI.** Detailed Information OF The 7 CI Participants in The Speech in Reverberation Test

| Subject | Gender | Age (yr) | CI Experience (yr) | CI Processor | Etiology |
| --- | --- | --- | --- | --- | --- |
| C14 | F | 41 | 12 | CP810 | Drug induced |
| C23 | M | 31 | 11 | CP810 | Sudden deafness |
| C30 | M | 13 | 10 | Freedom | LVAS |
| M5 | M | 18 | 15 | OPUS-2 | Virus infection |
| C16 | F | 25 | 2 | Freedom | Unknown |
| M17 | F | 18 | 5 | OPUS-2 | Genetic |
| M16 | F | 17 | 7 | OPUS-2 | Unknown |

The current results suggest that the first type GET vocoder is feasible to simulate speech perception with CIs, and the effects of CI current spread also could be simulated by manipulating durations of GETs. In both noise vocoder and GET vocoder, performance was substantially degraded by the increased current spread in both tasks. With eight or more bands, GET vocoders showed good simulation performance in that the actual CI data fell in the range between the separate and spread versions of the GETs.

## IV. Experiment 2. Simulation OF The N-of-M Strategy Ace

### A. Rationale

Some essential features of modern CI processing, including interleaved sampling, maxima selection, amplitude compression and quantization, are omitted in not only conventional continuous-carrier vocoders but also in the first type GET vocoder as used in Experiment 1. All of these features may influence speech perception. According to the analysis in Section II, GETs could be used to simulate them. The second type of GET vocoder [42, 43] is introduced here in detail, and a battery of speech recognition tasks was carried out to demonstrate its performance in Experiment 2. The experiment objective was to demonstrate the potential of CI speech perception simulation with a GET vocoder involving all of the above-mentioned essential features. The ACE strategy with 900-pps pulse rate was simulated by this advanced GET vocoder.

### B. Vocoder Theory: Direct mapping from electric pulses to GETs

In theory, the GETs are applicable for directly transferring any pulsatile CI electrodogram to a pulsatile vocoded sound. To be more illustrative, Fig. 7A demonstrates a 10-channel electrodogram (note: single vertical lines were used to represent electric pulses so that the amplitude and timing of the electric pulse can be represented, while the phase and gap durations in the common bi-phasic electric pulses were not considered in this study). To generate a GET vocoder, the 10 channels were converted into frequency bands spanning over 10 equally divided parts of the basilar membrane between characteristic frequencies of 150 and 8000 Hz [47]. The cutoff frequencies are 150, 271, 439, 672, 994, 1439, 2057, 2911, 4094, 5732, and 8000 Hz. Then, a band-specific GET was generated in this demonstration by setting the parameters in (1) as *a* = 1, *t*_0_ = 0, and

**Fig. 7.**
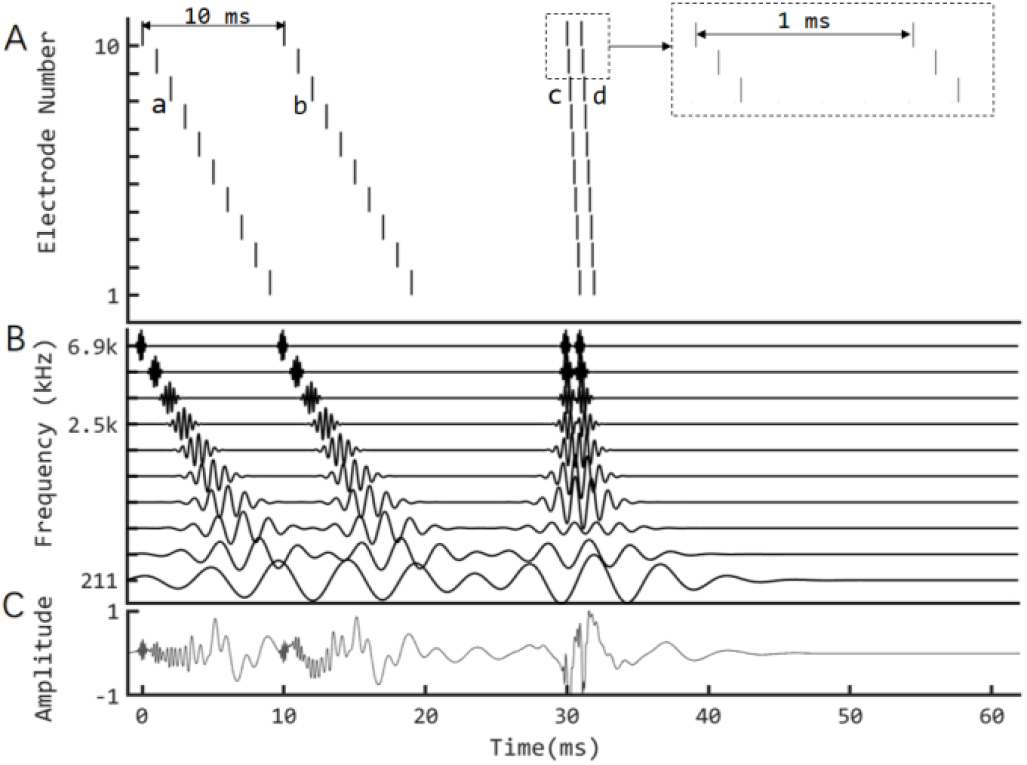
Mapping a CI electrodogram to a sound using the second type GET vocoder. A. An artificial 10-channel CI electrodogram, including two pulse sweeps with a 10-ms difference between a and b, as well as two additional sweeps with a 1-ms difference between c and d, corresponding to stimulation rates of 100 pps and 1000 pps, respectively. B. GETs mimicking the electric pulse trains. C. The final GET waveform resulting from the sum of ten band-specific GET trains in B.

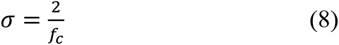

where *f*_*c*_ denotes the center frequency of the specific band. As a result, the band-specific GET had a 6.82-dB duration of

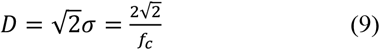

and a 6.82-dB bandwidth of

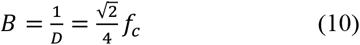

Then the acoustic GET train at the k^th^ channel in Fig. 7B is derived by

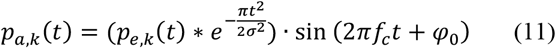

where *p*_*e,k*_(*t*) and *p*_*a,k*_(*t*) denotes the electric and acoustic pulse trains in Fig. 7A and 7B, respectively, “*∗*” denotes a convolution calculation, *σ* and *f*_*c*_ are band-dependent parameters as defined above, and *φ*_0_ is an initial phase that could be arbitrarily defined and was uniformly randomized between 0 and 2π here.

Fig. 7B shows the 10-channel GET trains, which have temporally separated waveforms for high-frequency channels, but overlapping waveforms for low-frequency channels. Fig. 7C shows the overall waveform summed from the 10 bands.

According to the theoretical analysis of GET simulation, pulsatile features for individual electric pulses cannot be guaranteed in the low-frequency channels, but the temporal-separation feature between groups of pulses may be simulated to some extent. For example, in Fig. 7B, at the lowest frequency channel, the 12-ms gap between b and c sweeps could have a counterpart, i.e., a shallow amplitude-modulation dip, in the waveform.

### C. Experiment method: Simulation of the n-of-m strategy ACE

Using the above method, any electrodograms, including the widely used *n*-of-*m* strategy like ACE strategy which is the current default strategy in Nucleus cochlear implants [45], can be converted to vocoded sounds. The specific vocoder is named ACE-GET. Following the preliminary results which showed comparable acute data between the ACE-GET vocoder and actual CI users [43], in this paper a battery of speech recognition tasks was carried out to further explore the potential of ACE-GET vocoder on simulation of speech perception with CIs.

In the clinical fitting of ACE strategy, the intensity dynamic range should be measured behaviorally electrode-by-electrode and is also limited and variable among users. In the ACE-GET vocoders, the dynamic range could be easily manipulated either in the compression stage of the ACE encoding or in the inverse compression stage of the GET synthesizing. The latter method was used in this study, and two dynamic ranges corresponding to two ACE-GET vocoders were tested. It was hypothesized that the vocoder with a higher dynamic range would simulate the top CI participants while the vocoder with a lower dynamic range would simulate the average performance of CI participants. The combination of *n* = 8 and *m* = 22 is one default option in the clinical fitting of ACE and was simulated in this experiment.

In detail, two 22-channel ACE-GET vocoders (denoted by GETlargeDR and GETsmallDR) were compared with two 22-channel sine-carrier conventional vocoders (125 Hz and 250 Hz envelope cutoffs, denoted by Sin250 and Sin125, respectively) with minimum channel overlapping as shown in Fig. 3A. The hypotheses for the parameter selection of the four vocoders are discussed later.

Detailed implementation methods of the *vocoders*: First, the default setting of the ACE software integrated in the CCi-Mobile software [49] was used to convert input sounds into electrodograms. An inverse-mapping function was used to transfer the electric current value of each electric pulse in the electrodogram to an envelope power value. Single-sample pulse trains from each band were “convolved” with a Gaussian function with *σ* = 3/*f*_*c*_. In the specific implementation of the experiment, the convolution step was replaced by simply comparing any overlapping sampling points from two GETs and preserving the larger point as the final sample value. In the theory and framework analysis in Section II, a convolution calculation was recommended, but in our experiment, we only preserved the largest point to show better pulsatile waveform than the cumulative effect of a convolution. The output was used to multiply a sinusoidal carrier with a frequency of *f*_*c*_ at the center of the corresponding band and an arbitrary initial phase (a random initial phase in this study). The average power of each band was kept unchanged. Finally, the modulated signals were summed to produce the vocoded stimulus.

The difference between GETlargeDR and GETsmallDR was only between their inverse (i.e., electric-to-acoustic) mapping functions, which are Eqs. 12 and 13, respectively:

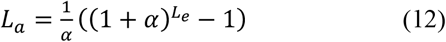

and

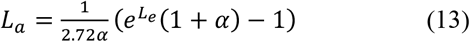

in which, the *L*_*a*_ denotes the recovered acoustic level, *L*_*e*_ denotes the electric current level defined by the electrodogram from the ACE strategy based on a specific patient’s fitting map, and *α* is a constant 416.0. In the present study, the threshold levels and most comfortable levels are constantly defined as 100 and 255 CU (current unit), i.e., 100 CU <*L*_*e*_ < 255 CU. In this case, based on (12) and (13), the recovered acoustic level ranges were 32.7 dB and 5.3 dB for GETlargeDR and GETsmallDR, respectively. The output stimuli level was controlled at a comfortable level around 65 dBA. Equation 12 is directly based on the default setting of the acoustic-to-electric compression function in ACE. It was hypothesized that GETlargeDR could simulate the best performance of CI listeners with the corresponding ACE strategy and GETsmallDR would significantly degrade the performance because of the much narrower range. Otherwise, the implementation details of the vocoder were the same as in [42].

In the two sine vocoders, the frequency spacing for cutoffs for the analysis filters was defined in the range of [80, 7999] Hz according to a Greenwood map [47]. Specifically, the cutoff frequencies were 80, 122, 172, 230, 298, 379, 473, 583, 712, 864, 1042, 1250, 1494, 1781, 2117, 2512, 2974, 3516, 4152, 4898, 5772, 6797, and 7999 Hz. The filtered signals were full-wave rectified and low-pass filtered (6th order Butterworth; 125 Hz for Sin125 and 250 Hz for Sin250) to extract the envelope for each channel. A sine wave with a frequency centered at the corresponding analysis band was used as the carrier, which was then multiplied by the corresponding envelope. The final vocoded stimuli were generated by a summation of the modulated carriers. In previous studies, it was found that speech intelligibility was better with a higher cutoff frequency in the envelope extraction [50]. Therefore, Sin250 was expected to be better than Sin125.

In Fig. 8, a Mandarin sentence was used to demonstrate the vocoded speech using the four vocoders, i.e., GETlargeDR, GETsmallDR, Sin250, and Sin125. It shows that the GET vocoders resemble the ACE-electrodogram more than the sine vocoders. The temporal separation between groups of pulses can also be found in the band signals of GET vocoded speech. Because the GET vocoders directly use the information of the ACE electrodogram, it was hypothesized that speech intelligibility would be worse, but closer to actual CI results, with the GET vocoders than with the sine vocoders.

**Fig. 8.**
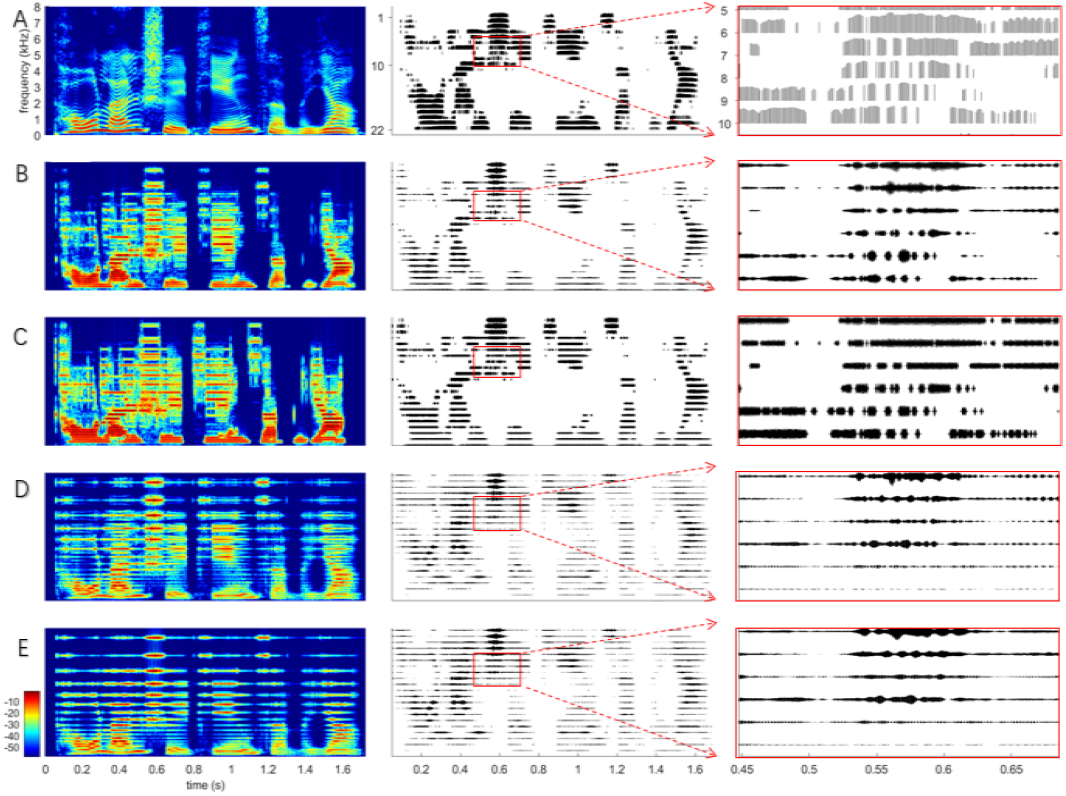
Speech stimulus demonstrations for the ACE-GET simulation experiment. Left: Spectrogram; middle: band-specific signal; right: zoom in of the boxed signals. A. Spectrogram and ACE electrodogram of a clear sentence of speech. B-E. Spectrogram and band-specific waveforms of vocoded speech using two GET vocoders (GETlargeDR, and GETsmallDR) and two conventional sine-wave vocoders (Sin250 and Sin125), respectively.

### D. Experiment method: Participants and Tasks

Two groups of NH participants (ten in each group, ages 18-29, and native Mandarin speakers) were tested in a soundproof room. Group 1 used Sin250 and GETlargeDR, and Group 2 used Sin125 and GETsmallDR. Three open-set Mandarin Chinese recognition tasks were tested, i.e., time-compression threshold, sentence-in-noise recognition, sentence-in-reverberation recognition. The results for the four tasks with the two vocoders in these NH participants were compared with actual CI results from our previous experiments [51] as well as newly collected data in this work. These experiments were conducted following procedures approved by the Medical Ethics Committee of Shenzhen University, China. Detailed information about the three experiments is as follows:

1. Time-compression thresholds (TCTs), i.e., accelerated sentence speeds at which 50% of words could be recognized correctly, were measured using the Mandarin speech perception corpus [52].
2. Speech reception thresholds (SRTs) in speech-shaped noise (SSN) and babble noise, i.e., signal-to-noise ratio (SNR) at which 50% of words could be recognized correctly, were measured using the Mandarin hearing in noise test (MHINT) corpus [53]. The TCT and SRT test procedures followed Experiment 2 of [51] strictly, in which ten CI subjects (9/10 adults) with various hearing histories were tested.
3. Recognition of speech in reverberation was measured using a Mandarin BKB-like sentence corpus [54], whose quiet sentences were convolved with simulated room impulse responses (RIRs). The RIRs were generated using a MATLAB function with its default setting, except the reverberation times (T60) were set as 0, 0.3, 0.6, and 0.9 s (https://www.audiolabs-erlangen.de/fau/professor/habets/software/rir-generator). For each T60, one sentence list was used. Seven CI participants with various hearing histories were also tested for comparison (See Table VI).

We had three subject groups, two of which were NH listeners each using two different vocoders. A mixed model was used to assess the repeated measures within subjects as well as independent measures between subjects. The paired-sample *t*-test and two-sample *t*-test were used to examine the statistical significance of the means’ difference for within-subject comparisons and between-subject comparisons, respectively. For each task, the five CI processing conditions, i.e., Sin250, Sin125, GETlargeDR, GETsmallDR, and CI, were pair-wisely examined to yield 10 pairs of comparison. Bonferroni corrections were used to adjust the p values, and the final significance was examined using the α level of 0.05.

### C. Results

The results with the four 22-channel vocoders, i.e., GETlargeDR, GETsmallDR, Sin250, and Sin125 are shown in Fig. 9.

**Fig. 9.**
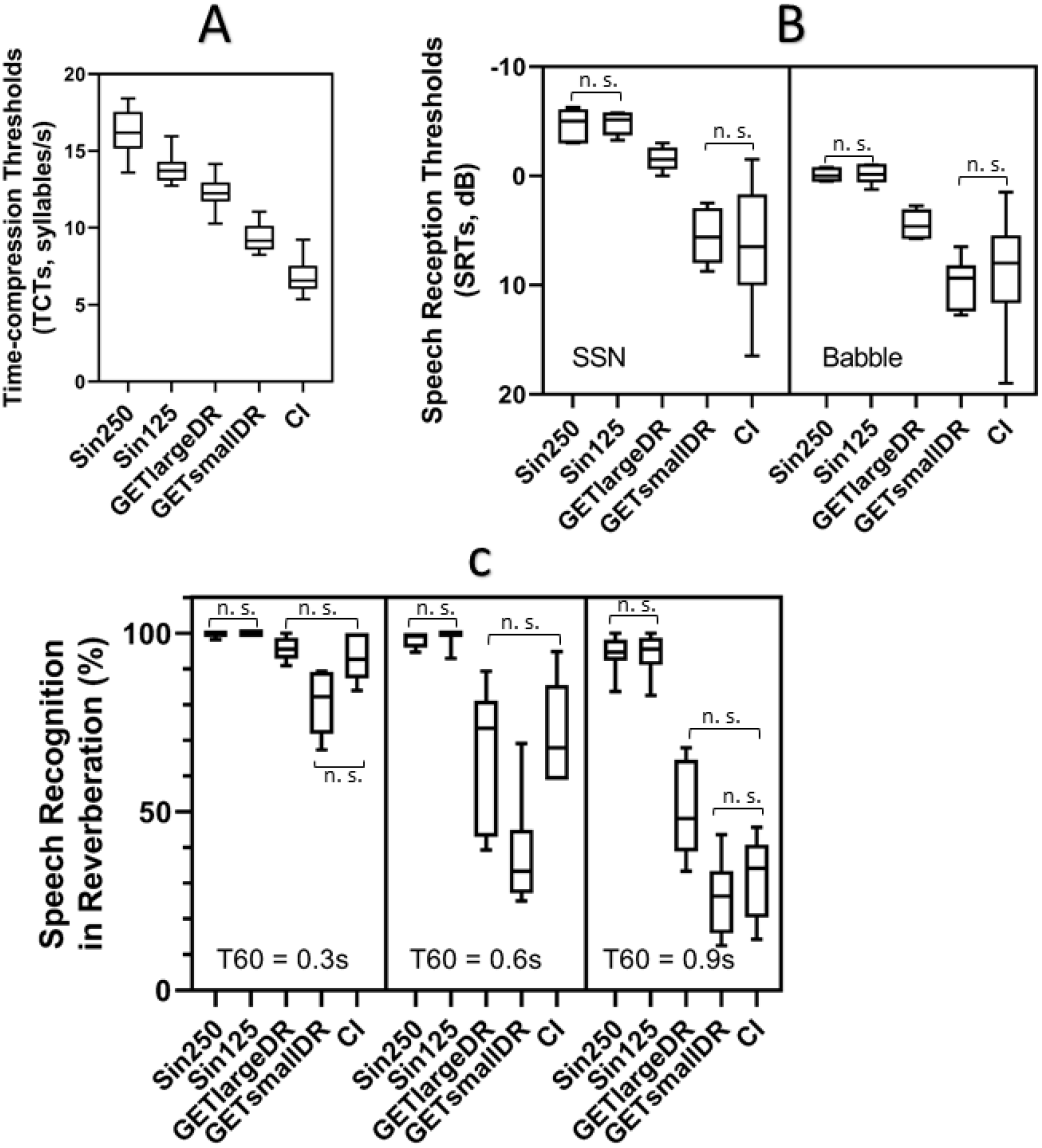
Results from three speech recognition tasks with two 22-channel sine-wave vocoders (Sin250: 250 Hz cut-off envelope; Sin125: 125 Hz cut-off envelope) and two GET vocoders (GETlargeDR and GETsmallDR; their difference is only in the intensity dynamic range, i.e., 32.7 dB and 5.3 dB for GETlargeDR and GETsmallDR respectively) compared with the results of some CI subjects. There were two groups of normal-hearing participants, each with ten participants. One group used Sin250 and GETlargeDR, and the other group used Sin125 and GETsmallDR. A. Time-compression threshold results. B. Speech reception threshold results of a speech in noise recognition experiment (SSN and babble noise). C. Speech recognition scores in reverberation with T60 = 0.3, 0.6, and 0.9s. Pairwise comparisons with Bonferroni corrections were examined. In each box, “n. s.” denotes the non-significant difference (*p* > 0.05), otherwise, there was a significant difference.

For the TCT test (Fig. 9A), a significant decreasing trend was found from Sin250 (mean = 16.1 syllables/sec), Sin125 (13.9), GETlargeDR (12.3), GETsmallDR (9.4), to actual CI (6.8) results (Bonferroni adjusted p < 0.05), while their standard deviations are comparable within the range from 1.0 to 1.2 syllables/s.

For the SRT test (Fig. 9B), there was no significant difference (adjusted *p* > 0.05) between Sin250 (means: −4.7 dB in SSN and −0.1 dB in babble noise) and Sin125 (means: −4.8 dB in SSN and −0.1 dB in babble noise) and between GETsmallDR (means: 5.6 dB in SSN and 10 dB in babble noise) and actual CIs (means: 6.5 dB in SSN and 8.8 dB in Babble noise). The mean results with GETlargeDR (means: −1.5 dB in SSN and 4.5 dB in babble noise) were significantly lower (adjusted *p* < 0.05) than those with Sin250 and Sin125, and significantly higher (adjusted *p* < 0.05) than those with GETsmallDR and CIs. The mean SRTs in babble noise were always significantly lower than those in SSN for all four vocoder conditions (adjusted *p* < 0.05). For CI users, mean SRTs in the two noise types did not show a significant difference (adjusted *p* > 0.05).

For the reverberant speech recognition test (Fig. 9C), all vocoders and the actual CI condition showed a significant trend of decreased recognition scores when the reverberation time increased. However, the NH listeners using sine vocoder simulations were much less sensitive to reverberation than the CI users. It is shown that even with T60 = 0.9 s, the sine vocoders still derived >94% means, which were much higher than CI participants’ 32%. The GETlargeDR and GETsmallDR derived significantly lower scores than the sine vocoders did (adjusted p < 0.05). Under the T60 = 0.3 s and 0.9 s conditions, there was no significant mean score difference between either GET vocoder and CI (adjusted *p* > 0.05), while GETsmallDR derived significantly lower mean scores than GETlargeDR did (adjusted *p* > 0.05). However, the mean results with CI were closer to GETlargeDR at T60 = 0.3 s and to GETsmallDR at T60 = 0.9 s. Under the T60 = 0.6 s condition, there was no significant mean score difference between GETlargeDR and CI, while GETsmallDR derived significantly higher mean scores than GETlargeDR and CI.

In all three tasks, GET vocoders resulted in more similar performance to actual CI performance than sine vocoders did. In fact, the sine vocoders overestimated CI performance in all tasks. Sin250 performed slightly better than Sin125 in mean results but did not show a significant difference. In the time-compression task, all vocoders produced better than CI performance, with GETsmallDR being the closest (Fig. 9A). In the SRT-in-noise test, GETsmallDR and CI produced comparable performance (Fig. 9B). In the reverberation task, GETlargeDR had similar-to-CI performance in all T60 conditions and GETsmallDR in the T60 = 0.3 and 0.9s conditions (Fig. 9C).

## V. Discussion

In this study, a GET-based vocoder was proposed, theoretically analyzed, and evaluated for its performance on CI speech perception simulation.

### A. GETs and electric pulses

The GET can be used to simulate a “perceivable” atom of sound, which can be traced back to Gabor (1947) [17]. More recently, it has been used in many psychoacoustic studies. The GET vocoder model can be a phenomenological one, in which each GET corresponds to an electrical pulse. The amplitude of the GET is scaled proportionally to the pulse current level. Moreover, the GET vocoders can simulate main features in CIs, including the place of stimulation, pulse time, temporal envelope, spectral envelope and spectral interaction, and intensity quantization and maxima-selection, by corresponding features of the acoustic pulses.

### B. Speech perception with GET vocoders

In this study, two types of GET vocoders (Fig. 3B & C) were proposed to simulate different aspects of CI processing [41, 42]. The first GET vocoder simply replaced the continuous noise or sine-wave carriers in conventional vocoders by a new type of carrier, or GET train. In the first implementation (Fig. 3B), a non-interleaved sampling 100-pps GET carrier was generated to study the effects of spread of excitation by controlling the GET duration according to the time-frequency uncertainty principle. Spread of excitation is an important factor underlying the poor- and large-variance performance for CI participants [44, 55-59]. Different from the noise-or sine-vocoders that produced performance better than actual CI performance even in the case of the severe channel interaction (i.e., using the low-pass filtered noise carriers), the GET vocoder produced a wide range of vowel and consonant recognition performance encompassing actual CI performance (Fig. 6). One limitation in this experiment was that the spectral spread simulated by GET vocoders at low frequency channels might be influenced by the sparsity of the electric pulses. For example (see Fig.7), at the lowest frequency channel, temporal overlap happens between two GETs and the bandwidth of the two overlapped GETs is narrower than an isolated GET. Fortunately, due to the sparse nature of speech signal and narrower GET durations at higher channels, the effects of this limitation should be limited. Another limitation of Experiment 1 was that all vocoders used a 50-Hz envelope cutoff frequency, which was lower than real CIs.

The second vocoder directly mapped individual electric pulses in a CI electrodogram to individual GETs to simulate the ACE strategy (Fig. 3C). This direct mapping allows simulation of all processing steps including the *n*-of-*m* maxima selection to amplitude compression and quantization. Compared with the conventional sine-wave vocoder, not only did the GET vocoder better resemble the ACE electrodogram, but more importantly the GET vocoder produced a mean and range of speech in noise recognition performance similar to that of actual CI users. In particular, the larger dynamic range simulated better CI performance (Fig. 9). Future studies are needed to establish and evaluate individualized CI simulation, in which both the mean and error patterns of phonemic recognition are used to judge the validity and quality of the simulation model [60-62].

The GET vocoder is perhaps a more general vocoder model as it can closely approximate conventional noise (using noise carriers instead of sine waves) and sine-wave vocoders by summing many GETs occurring at high rates or long GET duration and using high-fidelity intensity (or envelope) information. This means that the conventional vocoders can be treated as special cases of GET vocoders.

## VI. Conclusion

The main conclusions include:

1. The time-frequency uncertainty principle empowers and imposes constraints on using GETs for CI simulation;
2. Many features of modern CIs including pulsatile timing, current spread, n-of-m maxima selection, dynamic compression could be implemented in GET vocoders and then used to derive similar sentence recognition performance to actual CI users;
3. A GET vocoder framework for arbitrary CI strategy and a package of source code (using ACE as an example) are provided to serve as a general-purpose research tool to generate vocoded sounds (including speech) based on the direct pulse-to-pulse mapping.

Further experiment studies (e.g., in phoneme confusion patterns) are warranted to examine the performance of GET simulation systematically. The MATLAB source code of the GET vocoder for the ACE strategy is provided for academic research purposes (https://github.com/BetterCI/GETVocoder). Based on this code, more variants could be generated by manipulating the vocoder parameters, e.g., spectral spread, stimulation place or frequency shifting, and carrier types.

## Data Availability

All data produced in the present study are available upon reasonable request to the authors

https://github.com/BetterCI/GETVocoder

## Acknowledgment

We thank all the participants in these experiments. J. Carroll and S. Tiaden helped collect the data in Experiment 1. Fanhui Kong and Yulong Xiao helped collect the data in Experiment 2. Thanks to Drew Cappotto for proof-reading this article.

**Qinglin Meng** received the B.E. degree in electrical engineering from the Harbin Engineering University, Harbin, China, in 2008, and the Ph.D. degree in engineering from the Shanghai Acoustics Laboratory, Chinese Academy of Sciences, Beijing, China, in 2013. From 2013 to 2016, he was with Shenzhen University, Shenzhen, China, as a Postdoc. From 2017 to 2018, he visited the Department of Biomedical Sciences, City University of Hong Kong, Hong Kong, for one year. In 2016, he joined the Acoustics Laboratory, School of Physics and Optoelectronics, South China University of Technology, Guangzhou, China. His research interests include cochlear implant signal processing, sound perception, and hearing health.

**Huali Zhou** received the B.E. degree in Telecommunication Engineering from the Zhejiang University, Hangzhou, China, in 2007, and the M.S. degree in Acoustics from the South China University of Technology, Guangzhou, China, in 2021. From 2021, She is a Ph.D. student in the College of Electronics and Information Engineering, Shenzhen University, Shenzhen, China. Her research interests include sound signal processing, psychoacoustics, and cochlear implants.

**Thomas Lu**

**Fan-Gang Zeng** (S’88–M’91–SM’07–F’11) is the Director of the Center for Hearing Research and a Professor of anatomy and neurobiology, biomedical engineering, cognitive sciences, and otolaryngology—Head and neck surgery at the University of California, Irvine, CA, USA. He isa leading Researcher in hearing science and technology, with 288 publications, 16596 citations and an h-index of 662 (Google Scholar, June 28, 2022). He led development of the Nurotron26-electrode cochlear implant (SFDA approval in 2011 and CE Mark in 2012) and Sound Cure tinnitus suppressor (FDA clearance and CE Mark in 2011). He has consulted for NIH, NSF, DOD, and numerous other public and private agencies. He is the Editorial Board’s Chairman of the Hearing Journal and has been on the editorial board for six academic magazines. He served as the Chair of the2005 International Conference for Auditory Prostheses and edited three volumes on cochlear implants and tinnitus for Springer Handbook of Auditory Research. He holds 12 patents and has been on the Advisory Board and helped raise $80M for five medical device companies in U.S. and China.

## Notes

This research was supported by NIH R01 DC15587 (F.G.Z.), National Natural Science Foundation of China (11704129 and 61771320), Guangdong Basic and Applied Basic Research Foundation Grant (2020A1515010386), and Science and Technology Program of Guangzhou (202102020944) (Q.M.).

### Competing Interest Statement

The authors have declared no competing interest.

### Author Declarations

University of California Irvine Institutional Review Board and Medical Ethics Committee of Shenzhen University gave ethical approval for this work.

### Summary of Updates

Minor revisions have been made to improve the clarity.

